# Chronic pain and cognitive performance in dementia caregivers: Insights from the STELLA and DETECT-AD studies

**DOI:** 10.64898/2026.11.24.26368703

**Authors:** Christine McClure, Nora Mattek, Jeffrey Kaye, Walter D. Dawson, Allison Lindauer

## Abstract

**BACKGROUND:** Approximately 75% of individuals with Alzheimer’s disease and related dementias rely on informal caregivers, yet caregiver health, including chronic pain, remains under-addressed. Chronic pain is highly prevalent, functionally limiting, and linked to cognitive vulnerability, but remains understudied in informal caregivers. This study examined differences in pain and cognitive performance between caregivers and non-caregivers.

**OBJECTIVE:** To compare chronic pain and cognitive performance between dementia caregivers and non-caregivers and determine whether differences persist independent of age.

**METHODS:** We conducted a secondary analysis of longitudinal data from STELLA and DETECT-AD. Ninety-five participants were followed for 12 months. Weekly online questionnaires assessed pain intensity (0-10 scale) and pain interference (Likert scale), while annual in-person evaluations measured cognition using the Montreal Cognitive Assessment. Group differences were evaluated using t-tests and multivariate regression adjusting for age.

**RESULTS:** Groups were similar in sex and education but differed in age, pain, and cognition. Caregivers were significantly younger (71.3 vs 77.3 years; p < 0.0001) and caregivers demonstrated significantly higher global cognitive performance as measured by the MoCA (mean score 26.8 vs 25.2; p = 0.02). After age adjustment, caregivers reported significantly higher pain intensity (OR 1.42, 95% CI 1.00-2.01; p = 0.048).

**CONCLUSIONS:** Dementia caregivers exhibit greater pain burden despite younger age, highlighting chronic pain as a critical, underrecognized dimension of caregiver health and identifying an important opportunity to expand existing telehealth caregiver support models.

**CLINICAL TRIAL REGISTRY NAME, URL, AND REGISTRATION NUMBER:** STELLA: ClinicalTrials.gov, https://clinicaltrials.gov/study/NCT05338710, NCT04335110

DETECT: ClinicalTrials.gov, https://clinicaltrials.gov/study/NCT05385913

## BACKGROUND & OBJECTIVES

Approximately 75% of individuals living with Alzheimer’s disease and related dementias (ADRD) reside in community settings, where care is largely provided by informal, unpaid caregivers, most commonly spouses, partners, or other family members.^1,2^ This caregiving structure represents a foundational component of the dementia care ecosystem, enabling aging in place and reducing reliance on institutional care. At the same time, it transfers substantial responsibility to caregivers, whose health and functional capacity directly shape the trajectory of both their own well-being and that of the person living with dementia.

Over the past decade, there has been increasing recognition that sustaining community-based dementia care requires not only interventions for care recipients, but also scalable strategies to support caregivers.^3–12^ This shift is reflected in the rapid expansion of caregiver-focused interventions, particularly those delivered through telehealth platforms. Programs of technology-enabled adaptations of caregiver support interventions, including the “Support via Technology: Living and Learning with Advancing dementia” (STELLA)^11,12^ and Tele-Savvy, have demonstrated that remote delivery models are both feasible and acceptable for caregivers of people living with dementia.^3–18^ These approaches address key structural barriers to participation, including transportation, time constraints, and the logistical challenges of leaving a care recipient unattended, while allowing caregivers to engage in interventions within their home environments.

Evidence from these telehealth-based programs has consistently shown improvements in caregiver-centered outcomes, including reductions in perceived burden, depressive symptoms, and psychological distress, as well as gains in caregiving competence and self-efficacy. Collectively, this body of work has played a critical role in establishing telehealth as a viable and increasingly essential modality for caregiver support, particularly by demonstrating that comprehensive, multicomponent interventions can be successfully delivered in a scalable and accessible format.

Building on this strong foundation, there is increasing opportunity to extend these approaches by incorporating more targeted, symptom-specific strategies that address key drivers of caregiver health and functional capacity. While multicomponent interventions effectively address the broad and interconnected challenges of caregiving, certain high-impact symptoms may benefit from more focused attention within this framework. Among these, chronic pain emerges as a particularly important and underexamined factor, given its high prevalence, functional consequences, and potential contribution to both caregiver burden and downstream outcomes.^2,19–25^

Chronic pain is highly prevalent among caregivers of individuals with dementia, with estimates suggesting that more than half of caregivers experience ongoing pain.^2,20–24,26^ Pain in this population is associated with greater medication usage, more hospitalizations, and greater risk of chronic disease^19–21,24–25^ Importantly, caregiver pain has also been linked to adverse outcomes in care recipients and to broader system-level outcomes, including increased caregiver burden, higher likelihood of institutionalization for care recipients, and higher rates of care recipient mortality.^22,25,27^ Despite these associations, pain has received comparatively little attention within the caregiver intervention literature, particularly in the context of telehealth-delivered programs.

Caregivers are also at greater risk of memory loss and are up to six times more likely to develop dementia than non-caregivers.^28^ Pain and cognitive dysfunction are likely linked, as indicated by a growing body of research.^24–28^ Pain has been associated with impairments in attention, executive function, and memory, as well as with neurobiological changes involving stress pathways, inflammatory signaling, and neural network disruption.^29^ These mechanisms overlap with those implicated in both caregiving-related stress and neurodegenerative processes, suggesting that pain may represent a key pathway through which caregiving exposures translate into cognitive vulnerability.^30^

Despite these converging lines of evidence, the intersection of caregiving, chronic pain, and cognitive outcomes remains poorly characterized, and targeted interventions addressing pain in this population are notably lacking. To our knowledge, no published trials have specifically evaluated telehealth-delivered pain interventions in caregivers of people living with dementia. While telehealth interventions for chronic pain exist in other populations^30–32^ and caregiver teleinterventions targeting psychological outcomes are well-established,^3–18^ these two domains have not yet been meaningfully integrated.

The present work builds on this gap by leveraging data collected through STELLA^11,12^ and the Digital Evaluations and Technologies Enabling Clinical Translation for Alzheimer’s Disease (DETECT-AD)^33^ studies. STELLA provides a platform for engaging caregivers in telehealth-based support programming, while DETECT-AD enables longitudinal, real-world data collection through weekly online health questionnaires and annual cognitive assessments in individuals at risk for Alzheimer’s disease and their study partners. Together, these platforms create a unique opportunity to examine patterns of physical and cognitive health in caregivers within a naturalistic, technology-enabled context.

Rather than representing a prospectively designed trial focused on pain or cognition, the current analysis draws on existing longitudinal data collected as part of these programs to explore whether caregivers differ from non-caregivers in key domains of pain and cognitive performance. This approach allows for the identification of preliminary signals that may inform the design of future targeted interventions.

Extending these established caregiver support models to incorporate targeted, symptom-focused strategies represents a natural progression in the evolution of caregiver-focused care. Identifying and addressing high-impact, modifiable factors such as chronic pain within existing intervention frameworks may further enhance caregiver well-being and support early and preventative intervention to mitigate downstream health risks, while strengthening the sustainability of community-based dementia care.

## RESEARCH DESIGN & METHODS

All participants provided informed consent prior to study enrollment, and all study procedures were approved by the Oregon Health & Science University Institutional Review Board (DETECT: IRB# 23803 and STELLA: IRB# 19306). Participants were recruited from the Portland, Oregon metropolitan area through a combination of community-based advertisements, outreach presentations at retirement communities, and existing research registries affiliated with the Oregon Center for Aging and Technology (ORCATECH) and the Oregon Alzheimer’s Disease Research Center (OADRC).

Caregivers were enrolled in the Oregon Roybal Center for Care Support Translational Research Advantaged by Integrating Technology (ORCASTRAIT) parent study. Care partners then enrolled in STELLA as a pilot within ORCASTRAIT. Caregivers were defined as individuals providing ongoing, unpaid care to a person living with dementia. A comparison group of non-caregivers was selected from the DETECT-AD dataset, which contains data from participants in the same geographic region. Non-caregivers were selected to be demographically comparable with respect to key characteristics including sex and educational attainment. Participants were followed longitudinally for a period of 12 months.

### WEEKLY QUESTIONNAIRES

Pain-related outcomes were assessed using weekly online questionnaires distributed via the Qualtrics survey platform (Qualtrics, Seattle, WA).^34^ Participants received automated survey prompts at a consistent time each week. Pain intensity and pain interference were measured using validated visual analog scales. Pain intensity was rated on a scale from 0 (no pain) to 10 (worst possible pain), while pain interference was rated on a 5-point Likert scale ranging from 1 (not at all) to 5 (extremely), reflecting the extent to which pain interfered with daily activities.

Participants were instructed to report their pain experiences over the preceding week. Survey completion times were automatically time-stamped, allowing for precise tracking of adherence and response patterns. Additionally, participants entered event start and end dates using a calendar-based interface, enabling temporal alignment of reported symptoms. The specific questionnaire items used to assess pain intensity and interference are presented in Table 1. To distinguish persistent pain from intermittent or isolated pain, weekly pain reports were reviewed longitudinally to identify participants reporting pain over a period of at least three months, consistent with the temporal definition of chronic pain.

**TABLE 1.** PAIN INTENSITY AND PAIN INTERFERENCE QUESTIONS QUERIED WEEKLY.

|  |  |
|---|---|
| PAIN LEVEL | Please rate your pain by indicating the number (0-10) that best describes your pain on average. |
| PAIN INTERFERENCE | During the past week, how much did pain interfere with your normal activities or work (1-5)? |

\*For pain level, 0 indicates no/lowest pain and 10 indicates highest pain level. For pain interference, 1 indicates lowest pain interference and 5 indicates highest pain interference.

### COGNITIVE ASSESSMENTS

All participants underwent annual clinical evaluations coordinated through ORCATECH. These assessments included the National Alzheimer’s Coordinating Center (NACC) Uniform Data Set (UDS, Version 3), which comprises a standardized neuropsychological battery designed to assess multiple cognitive domains. Global cognitive function was evaluated using the Montreal Cognitive Assessment (MoCA),^35^ a widely used screening tool for mild cognitive impairment and early dementia. Some participants completed the 22-point Blind MoCA, which excludes items requiring visual input. These scores were converted to equivalent 30-point MoCA scores using published equipercentile score equivalencies^36^ to facilitate comparison across participants. Individuals with a diagnosis of dementia were excluded from participation in both DETECT-AD (controls) and STELLA (caregivers).

Comorbid medical conditions were assessed at baseline using the Cumulative Illness Rating Scale (CIRS),^37^ providing a standardized measure of overall disease burden. All evaluations were conducted by trained clinicians using standardized protocols to ensure consistency and reliability across participants.

### STATISTICAL ANALYSES

Summary statistics (mean, standard deviation (SD) or count [%]) for demographics and baseline clinical measures were generated for caregivers and non-caregivers. Differences in caregivers and non-caregiver’s participant characteristics were statistically compared using 2-sample independent t tests or Wilcoxon rank sum tests as appropriate for continuous variables and Fisher’s exact test or chi-square test as appropriate for categorical variables. Mean pain intensity and pain interference levels were calculated for each participant over the one-year period, and these were compared between groups using 2-sample independent t-tests. Multivariate logistic regression was used to examine the associations between pain metrics and caregiver/control groups after adjusting for age. Analyses were performed using SAS software (version 9.4, SAS Institute).

## RESULTS

The analytic sample consisted of 95 participants, including 21 caregivers (from the STELLA cohort) and 74 non-caregivers (from the DETECT cohort). Participant demographic information is listed in Table 2.

**TABLE 2:**
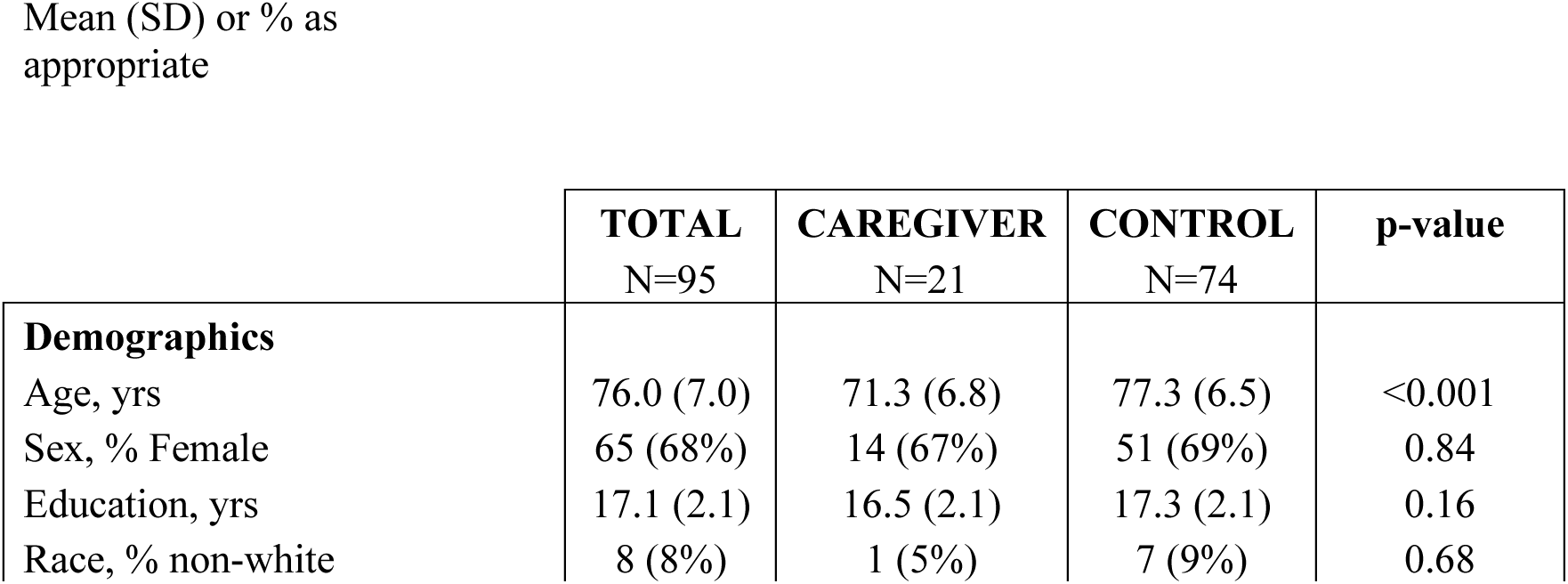
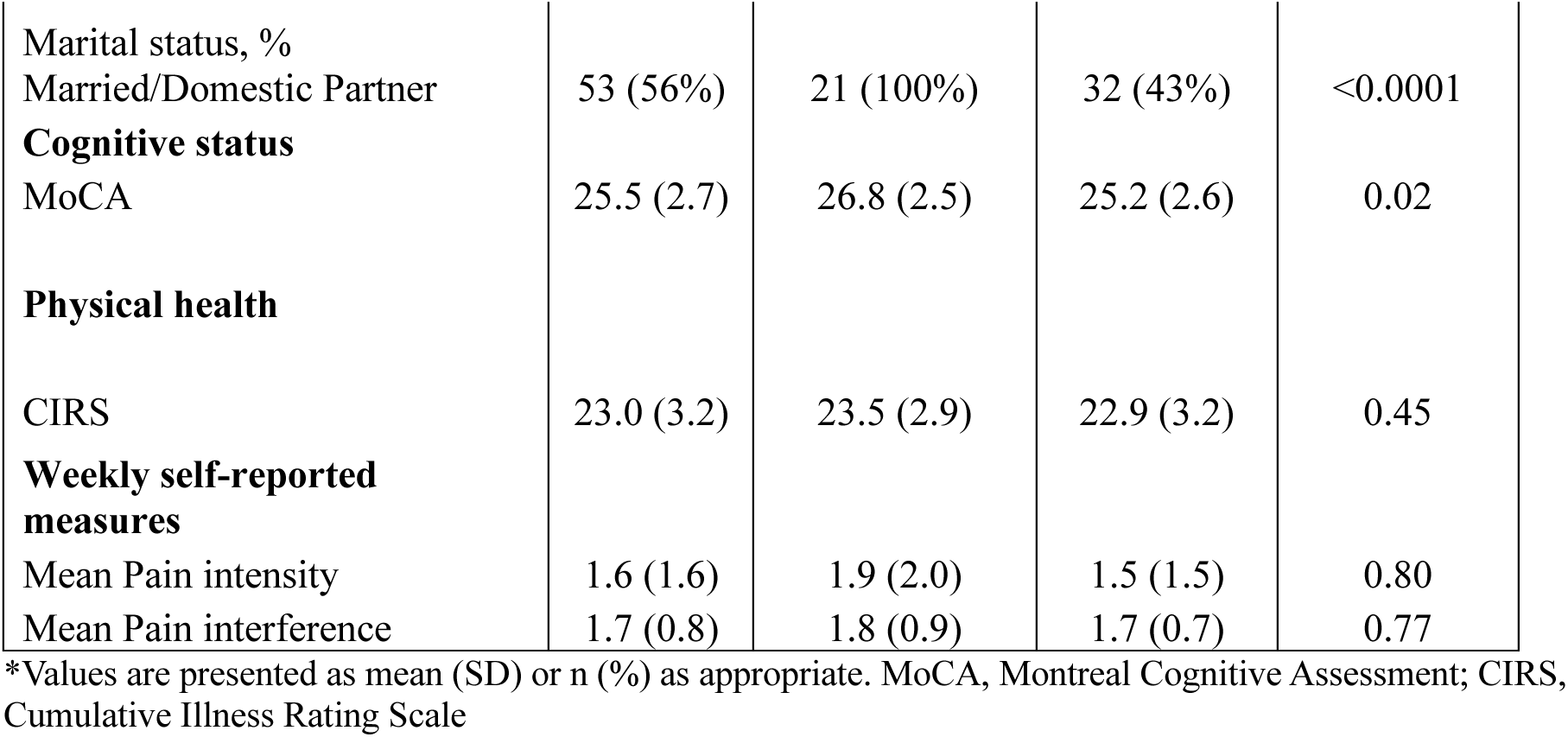
CAREGIVER AND CONTROL PARTICIPANT CHARACTERISTICS.

|  | **TOTAL**<br>N=95 | **CAREGIVER**<br>N=21 | **CONTROL**<br>N=74 | **p-value** |
|---|---|---|---|---|
| **Demographics** |  |  |  |  |
| Age, yrs | 76.0 (7.0) | 71.3 (6.8) | 77.3 (6.5) | <0.001 |
| Sex, % Female | 65 (68%) | 14 (67%) | 51 (69%) | 0.84 |
| Education, yrs | 17.1 (2.1) | 16.5 (2.1) | 17.3 (2.1) | 0.16 |
| Race, % non-white | 8 (8%) | 1 (5%) | 7 (9%) | 0.68 |
| Marital status, % Married/Domestic Partner | 53 (56%) | 21 (100%) | 32 (43%) | <0.0001 |
| **Cognitive status** |  |  |  |  |
| MoCA | 25.5 (2.7) | 26.8 (2.5) | 25.2 (2.6) | 0.02 |
| **Physical health** |  |  |  |  |
| CIRS | 23.0 (3.2) | 23.5 (2.9) | 22.9 (3.2) | 0.45 |
| **Weekly self-reported measures** |  |  |  |  |
| Mean Pain intensity | 1.6 (1.6) | 1.9 (2.0) | 1.5 (1.5) | 0.80 |
| Mean Pain interference | 1.7 (0.8) | 1.8 (0.9) | 1.7 (0.7) | 0.77 |

\*Values are presented as mean (SD) or n (%) as appropriate. MoCA, Montreal Cognitive Assessment; CIRS, Cumulative Illness Rating Scale

Caregivers and non-caregivers did not differ significantly with respect to sex distribution or educational attainment, suggesting successful demographic matching across these variables.

However, significant between-group differences were observed in age and cognitive performance. Caregivers were significantly younger than non-caregivers (mean age 71.3 vs 77.3 years; p < 0.0001) and caregivers demonstrated significantly higher global cognitive performance as measured by the MoCA (mean score 26.8 vs 25.2; p = 0.02; Figure 1). The low MoCA scores observed among several non-caregiver participants did not reflect dementia diagnoses; three of these participants were clinically classified as having MCI and one as cognitively normal.

**FIGURE 1:**
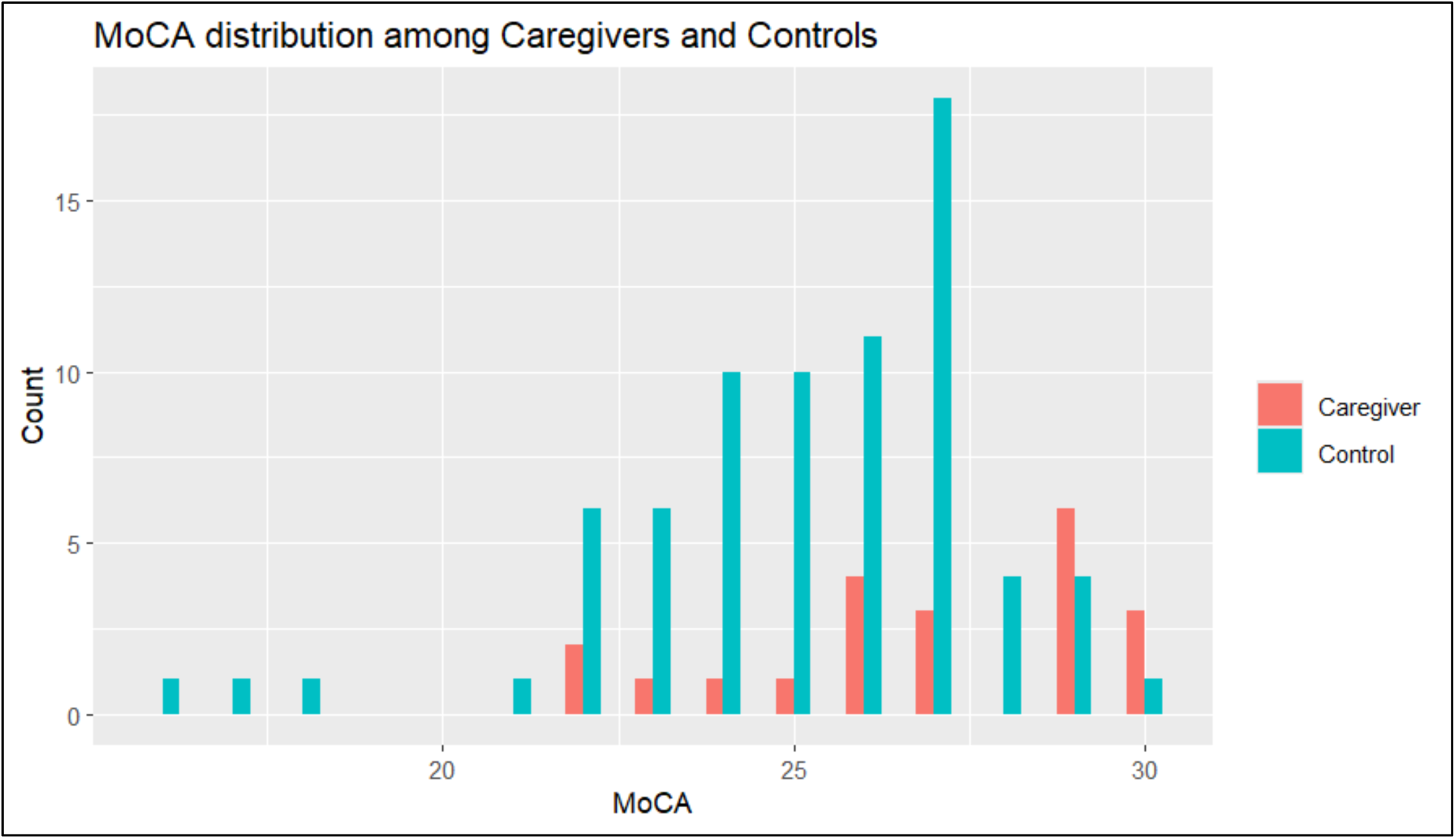
MoCA SCORES

With respect to pain outcomes, after adjusting for age caregivers reported significantly greater pain intensity compared with non-caregivers over the 12-month follow-up period (odds ratio = 1.42, 95% confidence interval = 1.00–2.01; p = 0.048).

**FIGURE 2:**
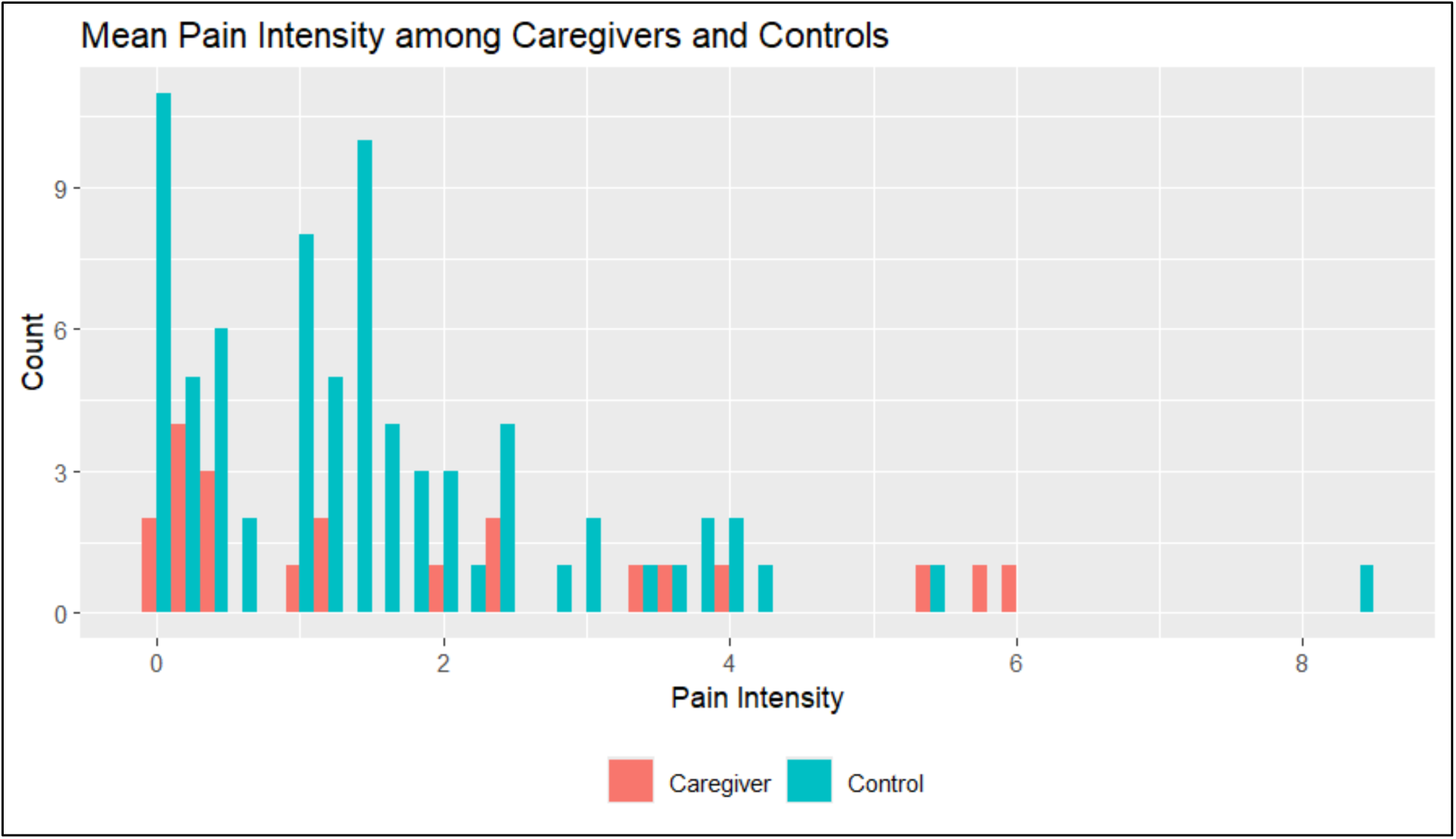
PAIN INTENSITY SCORES

After adjusting for age, caregivers reported greater pain interference compared with non-caregivers over the 12-month follow-up period (odds ratio = 1.89, 95% confidence interval = 0.89 – 4.02; p = 0.096).

**FIGURE 3:**
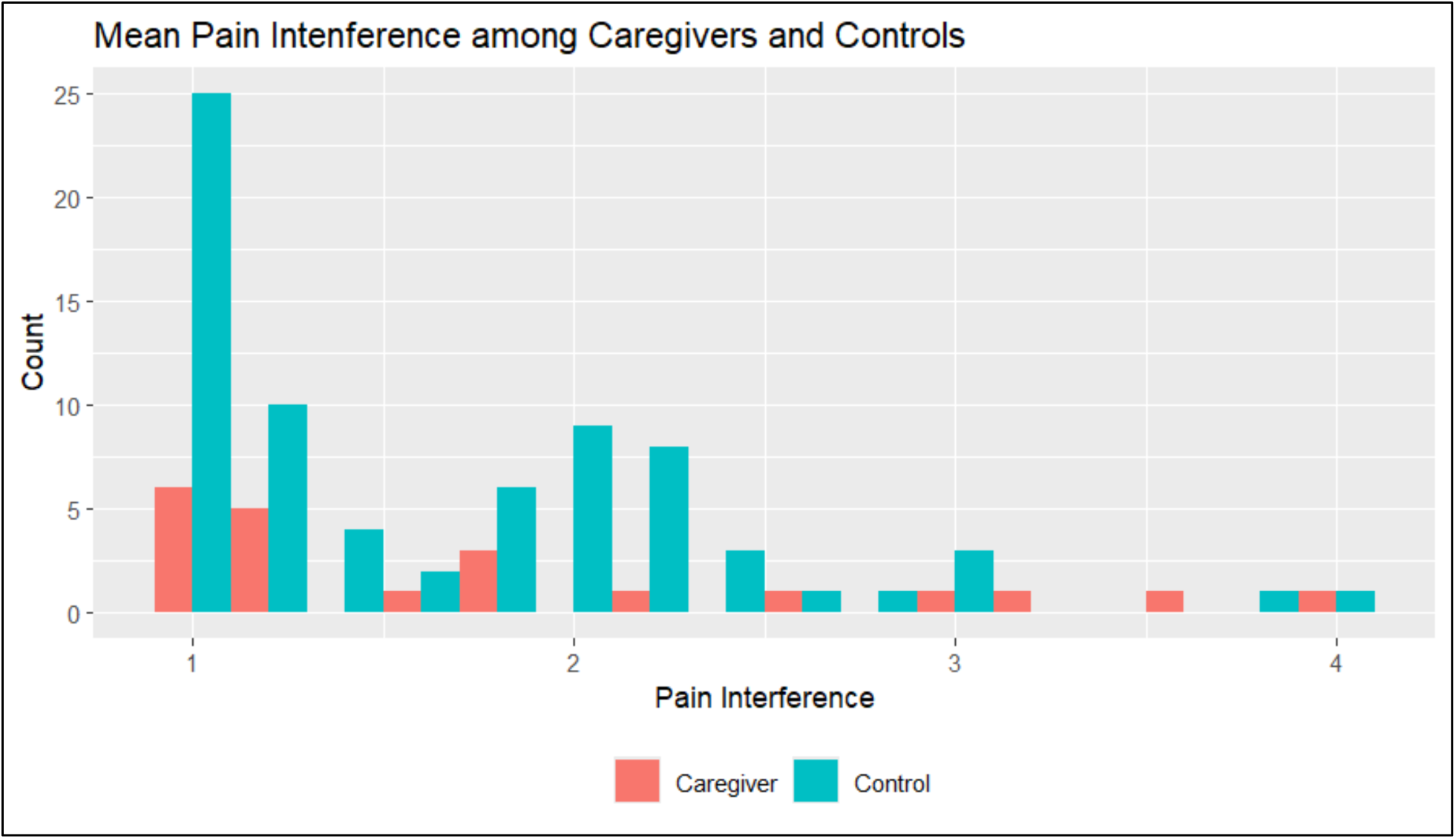
PAIN INTERFERENCE SCORES

Although average pain severity was relatively low, 12 of 21 caregivers (57.1%) and 50 of 74 non-caregivers (67.6%) reported pain persisting for at least three months, consistent with the temporal definition of chronic pain.

## DISCUSSION & IMPLICATIONS

Leveraging data collected weekly through STELLA and DETECT-AD longitudinal monitoring infrastructure, we found that caregivers of individuals living with dementia experienced higher pain burden and higher cognitive performance compared with non-caregivers. Notably, these differences were observed despite caregivers being younger on average, suggesting that the relationship between caregiving, pain, and cognition may be more complex than previously recognized and warrants further investigation.

Importantly, these findings emerge not from a study originally designed to examine pain or cognition, but from the secondary analysis of data collected within telehealth-enabled caregiver support and monitoring platforms. As such, they highlight the value of leveraging existing data streams to identify underrecognized patterns of risk and to inform the next generation of targeted interventions.

Within the broader landscape of caregiver intervention research, these results highlight an important opportunity for further development. Telehealth-delivered programs such as Tele-Savvy and STELLA have demonstrated clear benefits in reducing caregiver stress, burden, and depressive symptoms, establishing a strong foundation of effective, multicomponent support strategies^3–18^ Building on this foundation, there is growing opportunity to extend these approaches by incorporating attention to specific, high-impact symptoms that may influence caregiver health trajectories and downstream outcomes.

Chronic pain represents one such symptom. It is highly prevalent, functionally limiting, and strongly associated with caregiver burden, quality of life, and care recipient outcomes. At the same time, it is potentially modifiable and amenable to intervention.^20,22^ Despite this, there remains a notable absence of telehealth-delivered pain interventions specifically designed for caregivers of individuals living with dementia. This gap is particularly striking given the alignment between telehealth delivery models and the practical needs of caregivers, as well as the growing evidence supporting telehealth approaches in both caregiver support and chronic pain management separately.

The findings of the present analysis support the development of targeted, telehealth-delivered pain interventions for caregivers as a complementary extension of existing approaches. Such interventions would build on the established feasibility and acceptability of telehealth in this population while incorporating attention to an important and underexamined determinant of caregiver health. Integrating a more focused emphasis on pain alongside broader support strategies may further enhance functional capacity and daily caregiving ability.

From a translational perspective, this work provides a foundation for progressing from observational insights toward interventional development within established telehealth frameworks. The integration of telehealth delivery with longitudinal monitoring platforms offers an opportunity to design adaptive, scalable interventions that can be embedded within existing caregiver support infrastructures. Targeting pain as a modifiable factor may also have cascading benefits, not only for caregivers but also for care recipients, by enhancing caregiving capacity and potentially delaying institutionalization.

Several limitations should be acknowledged. As an exploratory analysis of existing data, causal relationships cannot be established, and the sample size is modest, recruited from one geographically specific region. Additionally, the caregiver and non-caregiver groups differed in age, although analyses adjusted for this factor. Importantly, overall pain severity in this cohort was relatively low, and participants were not recruited on the basis of pain. This may also represent a strength of the present analysis, as the findings reflect pain experienced within a community-based sample rather than a population selected for high pain burden. Despite relatively low average pain intensity, more than half of caregivers (57.1%) and nearly two-thirds of the overall sample (65.3%) reported pain persisting for at least three months, consistent with the temporal definition of chronic pain. This finding is broadly consistent with prior estimates indicating that chronic pain affects more than half of older adults.^2,20–24,26,38–46^ Thus, low average pain severity did not necessarily reflect isolated or occasional pain.

Future work should build on these findings through the development and testing of targeted telehealth pain interventions for caregivers of individuals living with dementia, particularly among caregivers with persistent or clinically significant chronic pain. Randomized controlled trials will be needed to determine whether reducing pain in caregivers leads to improvements in functional, psychological, and cognitive outcomes. Mechanistic studies incorporating physiological and behavioral measures may further elucidate the pathways linking pain, caregiving, and cognitive function, and help identify which caregivers are most likely to benefit from targeted pain interventions.

In conclusion, this study highlights chronic pain as a critical, underrecognized dimension of caregiver health and identifies an important opportunity to expand existing telehealth caregiver support models. By demonstrating preliminary associations among caregiving, pain, and cognitive function, these findings provide a strong foundation for future research investigating the complex relationships among these factors and for the development of focused, scalable interventions that incorporate targeted pain management within established telehealth frameworks. Addressing caregiver pain through telehealth may represent a promising strategy for improving caregiver well-being, preserving caregiving capacity, and strengthening the sustainability of community-based dementia care.

## Acknowledgements

The authors would like to sincerely thank all study participants for their time, commitment, and willingness to contribute to this research. Their participation made this work possible and represents an invaluable contribution to advancing our understanding of caregiver health and well-being. The authors also acknowledge the excellent assessment and follow-up provided by faculty and staff of the STELLA and DETECT-AD.

## Author Contributions

Conception or design of the work: C. McClure, J. Kaye, A. Lindauer. Acquisition of data: W.D. Dawson Analysis of data: N. Mattek. Interpretation of data: C. McClure, N. Mattek, J. Kaye, A. Lindauer. Drafting of the manuscript: C. McClure, N. Mattek Revision of the manuscript: C. McClure, N. Mattek, J. Kaye, A. Lindauer. All authors approved the final version to be published.

## STATEMENTS AND DECLARATIONS

### Ethical Considerations

The study protocol received ethical approval from the OHSU Institutional Review Board (IRB # 19306).

### Consent to Participate

All participants provided written informed consent prior to participating in this study.

### Consent for Publication

Not applicable.

### Data Availability

The data supporting the findings of this study are available from the Oregon Life Laboratory within the Digital Technology Core (DTC) of the Oregon Alzheimer’s Disease Research Center (OADRC) and may be requested at: https://octri.ohsu.edu/redcap/surveys/?s=EW4HKW47XH&_ga=2.9914869.1166959164.1653610093-1864845509.1632940067

